# Impact of Body Composition on Peak Oxygen Uptake After Cardiac Rehabilitation

**DOI:** 10.1101/2025.08.22.25334270

**Authors:** Wiaam Elkhatib, Thomas Olson

## Abstract

**Background:** Peak oxygen uptake (VO_2_ peak), standardized to total body mass (ml/kg/min) is a predictor of morbidity and mortality in cardiovascular disease patients. However, subsets of individuals undergoing exercise based cardiac rehabilitation (CR) show no improvement or reduction in VO_2_ peak despite improvement in other functional measures. Our aim is to assess the influence of CR on lean mass, and the effect of body composition compared to total body mass changes on peak VO_2_ following CR.

**Methods:** This pre-post intervention study included adults >18 years old who completed CR between 2015-2022 at Mayo Clinic, Rochester. All patients completed both dual energy X-ray absorptiometry (DXA) and cardiopulmonary exercise testing (CPET) for measurement of body composition (total mass and lean mass) and VO_2_ peak pre-post CR. Improvement in VO_2_ peak was defined as positive percent change. Descriptive statistics, paired t-tests, univariable and multivariable linear regression modeling were performed.

**Results:** Of 140 subjects, 19.3% were female and 96.4% White with a mean (SD) age of 63 (12.5) years, BMI of 30.2 (5.82), and a mean of 27.1 (11.7) completed number CR sessions. Pre-post CR total body mass loss was −1.28 (3.18) kg and lean mass gain was 0.84 (2.86) kg. All changes were statistically significant (p<0.05). Pre-post CR VO_2_ peak in absolute units had a percent change increase of 6.52 (13.1) mL/min, relative (corrected for total body mass) increase of 8.24 (13.6) mL/kg/min, and relative to lean mass increase of 5.73 (13.4) mL/lean-kg/min. Pre-post percentage of subjects with a positive change in peak VO_2_ in absolute units was 70%, relative was 75%, and relative to lean mass was 67.9%. Multivariable regression showed statistical significance in peak VO_2_ percent change for all units when adjusted for pre-CR peak VO_2_.

**Conclusions:** Our findings demonstrate significant increases in lean mass and VO_2_ peak following CR, with larger improvements reflected in units adjusted for total mass compared to other methods. These data suggest the reporting methodology for change in VO_2_ peak following CR impact overall results.

**CLINICAL PERSPECTIVE:**

- Published studies have reported that subsets of patients undergoing exercise-based cardiac rehabilitation (CR) may show no improvement or even reduction in peak oxygen uptake (VO_2_ peak), a predictor of mortality in individuals with cardiovascular disease, which may be partly influenced by body composition.
- This pre-post intervention study of 140 adult patients who completed CR demonstrated significant increases in lean mass and VO_2_ peak following CR across the cohort while suggesting the reporting methodology for change in VO_2_ peak impacts overall results.
- Our findings could imply that a proportion of patients continue to demonstrate additional abnormalities that inhibit improvement in VO_2_ peak independent of body composition alterations, partially explaining why a subset of patients have a lack of positive change in VO_2_ peak.
- Significant predictors for reduced VO_2_ peak improvement following CR may include female sex and pre-CR VO_2_ peak.
- Despite CR significantly improving lean body mass, the ability to utilize that mass to generate useful oxidative metabolism could be limited and may require a longer duration of CR to maximize oxidative metabolic efficiency and improve patient outcomes following acute cardiac events.

## INTRODUCTION

Measurement of peak oxygen uptake (VO_2_ peak) is considered the gold standard metric of cardiopulmonary fitness and is inversely related to hospitalization and mortality, independent of age, sex, race, and comorbid conditions [1]. This metric is most used clinically, via cardiopulmonary exercise testing (CPET), to determine cardiorespiratory fitness, examine origin of exertional symptoms, as a surveillance tool for prognosis and timing of interventions, and determine efficacy of interventions. Importantly, as VO_2_ peak is inversely related to hospitalizations and mortality, clinical interventions designed to improve VO_2_ peak are imperative. Exercise based cardiac rehabilitation (CR) is a Class 1 therapy for the secondary prevention of cardiovascular disease [2]. Exercise training, as prescribed during CR, is known to have a positive impact on modifiable cardiovascular disease risk factors (e.g. obesity, diabetes, hypertension, etc.) including improving VO_2_ peak. Interestingly, a proportion of CR patients are known to have no improvement, or even a reduction, in VO_2_ peak after CR [3].

The absolute volume of oxygen an individual can consume is related to the size/mass of the individual. Therefore, larger individuals will, by definition, consume larger amounts of oxygen to maintain vital organ function. As such, the units to describe VO_2_ peak are often standardized to total body mass to make comparisons across populations in which body mass differs, allowing for more accurate comparisons across different patient groups [4]. However, bias using these units may arise towards obese patients (with larger, less metabolically active adipose stores) relative to other subsets of patients (with larger amounts of highly metabolically active lean muscle mass) since the composition of the total body mass is not accounted for [5]. With this, overweight persons with high body fat demonstrate a lower VO_2_ peak relative to their body mass, though difficult to differentiate from persons with high lean body mass [6]. This may directly influence the results of interventions specifically designed to improve VO_2_ peak, body composition, or both, as may be the case with CR. Therefore, the change in VO_2_ peak based on total mass may not reflect true change in fitness for patients who differentially alter body composition (e.g., increase lean mass vs. decrease fat mass) following CR.

Studies examining the influence of body composition on aerobic performance in cardiac rehabilitation are limited despite VO_2_ peak normalization serving as a determining factor in results interpretation [7]. Assessment of change in VO_2_ peak based on total body mass alone may not be optimal. Therefore, the objective of this study was to assess the influence of CR on lean mass and the effect of body composition compared to total body mass changes on VO_2_ peak following CR. We hypothesized that the change in VO_2_ peak post-CR, adjusted for lean mass rather than total mass, will better reflect improvements in VO_2_ peak.

## METHODS

### Study Design

Reporting of data adheres to STROBE guidelines [8]. This retrospective pre-post intervention cohort study at a large tertiary-care center which included consecutive clinically stable adult patients over the age of 18 years referred to and enrolled in an early outpatient (phase II) comprehensive exercise-based CR program between January 2015 to October 2022. The CR occurred at the Mayo Clinic Cardiovascular Health Center in Rochester, Minnesota within 2-to-4 weeks following referral after major cardiac event fulfilling approved clinical indication for CR. All included patients approved to have their medical records used for research purposes. The Mayo Clinic Institutional Review Board approved this study. Involved patients had provided medical record use authorization.

### Patient Identification

Clinical and demographic characteristics were electronically obtained using resources of the Rochester Epidemiology Project (REP), a records linkage system that captures data on health conditions (diagnosis, procedures, and other vital information) of all Olmsted County residents who receive medical care at Olmsted Medical Center or the Mayo Clinic and its allied health care centers.

### Cardiac Rehabilitation

The Mayo Clinic comprehensive outpatient CR program adheres to the American Association of Cardiovascular and Pulmonary Rehabilitation established guidelines [9]. A total of 36 supervised CR sessions were prescribed over a 3 days per week regimen completed over 12 weeks. Sessions involved supervised aerobic activity for 20-45 minutes followed by resistance training for 10-15 minutes. Equivalent home-based aerobic exercise was recommended on days outside of supervised CR sessions. Individualized exercise prescriptions utilizing the Borg rating of perceived exertion scale were updated as patients advanced through their program, with all patients having attended a minimum of 1 documented session [10]. Ancillary lifestyle education was provided and centered around sleep hygiene, nutrition, medication adherence, smoking cessation (as necessary), and cardiovascular risk factors modification.

### Data Collection

Patients had multiple indications recorded for CR program referral obtained via documental medical record review. Sex and birth date were logged. Height and weight measures were documented for computation of body mass index (BMI, kg/m2).

Symptom-limited maximal cardiopulmonary exercise testing (CPET) via motor-driven treadmill pre-post CR was obtained electronically from an institutional registry and used to ascertain VO_2_ peak. Medications were not withheld prior. When multiple tests were identified, the test performed closest to CR enrollment and graduation were used for analysis. The CPETs incorporated an institutionally designed incremental exercise protocol increasing 2 metabolic equivalents every 2 minutes under cardiologist oversight by a specially trained nurse or exercise physiologist [11]. At each stage, the rating of perceived exertion and manual sphygmomanometry were completed alongside a continuous 12-lead electrocardiogram for safety monitoring. Indirect calorimetry was used to measure gas exchange variables (MGC Diagnostics). Peak values were recorded for oxygen consumption, carbon dioxide production, O_2_ pulse (oxygen consumption divided by heart rate) and respiratory exchange ratio averaging over 10 seconds during the test. All patients included in this analysis achieved a peak respiratory exchange ratio of equal to or greater than 1.0 during both pre-post CR CPETs completed within 12 months of program graduation [12].

All pre-CR dual energy X-ray absorptiometry (DXA) was completed after their CR qualifying event and prior to enrollment in CR whereas the post-CR DXA was completed within 4 weeks of graduation (GE Lunar iDXA whole body scanner). Standard institutional procedure was followed. Relevant body composition measurements included total body mass (kg) and lean mass (kg).

VO_2_ peak units were reported using CPET and DXA results in three different units. Absolute units (mL/min) constituted the first method of reporting. Units relative to total body mass were reported as absolute units divided by total body mass (mL/kg/min). Units relative to lean mass were reported as absolute units divided by lean body mass (ml/lean-kg/min).

### Statistical Analysis

Categorical variables were descriptively reported as number (percentage) and continuous variables were reported as mean values with standard deviation (SD). Improvement in VO_2_ peak for each of the three reported unit groups before and after CR was defined as a positive ( >0) percent change and/or positive mean difference. Paired student t-tests were completed for pre-post CR measures of VO_2_ peak per unit, total body mass and lean body mass. For the primary outcome, linear regression models for the percent change in VO_2_ peak per reported unit group were constructed to compare the VO_2_ peak percent change of the cohort to the population at large after controlling for age, sex, pre-CR BMI, number of CR sessions, and pre-CR VO_2_ peak per unit. Beta coefficients were provided with standard errors (SE), 95% confidence intervals (CI), and p-values. Statistical significance for all methods was defined as a *p-value* <0.05. Variable selection was determined by clinical expertise, univariable analysis, and previous literature. All statistical analyses were conducted via RStudio Version 4.4.2 (RStudio, PBC).

## RESULTS

Of 140 patients, 19.3% were female. Caucasian patients constituted 96.4% of the cohort. The mean age was 63.0±12.5 years. Mean pre-CR BMI was 30.2±5.8. On average, patients completed 27.1±11.7 supervised CR sessions. The most common CR indications were percutaneous coronary intervention [81 (58.7)] and acute coronary syndrome [66 (47.8)]. Demographics and CR indication are provided in Table 1.

**Table 1:** Patient demographics are outlined by frequency [n (%)] or mean (SD) as appropriate. CR = cardiac rehabilitation; BMI = body mass index; ACS = acute coronary syndrome; CABG = coronary artery bypass graft; PCI = percutaneous coronary intervention.

| Patient Demographics [n=140] |  |  |
| --- | --- | --- |
| Characteristics |  | Totals |
| Female Sex |  | 27 (19.3) |
| Age (years) |  | 63 (12.5) |
| Number CR Sessions |  | 27.1 (11.7) |
| Pre-CR BMI (kg/m<sup>2</sup>) |  | 30.2 (5.8) |
| Race | Caucasian | 135 (96.4) |
|  | African American | 1 (0.7) |
|  | Other/Unknown | 4 (2.9) |
| CR indication | ACS | 66 (47.8) |
|  | Angina | 17 (12.3) |
|  | Heart Failure | 5 (3.6) |
|  | CABG | 6 (4.3) |
|  | PCI | 81 (58.7) |
|  | Valvular Surgery | 8 (5.8) |
|  | Heart Transplant | 2 (1.4) |
|  | Other | 2 (1.4) |

Pre-post CR VO_2_ peak in absolute units (mL/min) had a percent change increase of 6.5±13.1%, relative to total body mass units (mL/kg/min) increase of 8.2±13.6, and relative to lean mass units (mL/lean-kg/min) increase of 5.7±13.4. Pre-post CR delta for VO_2_ peak in absolute units (mL/min) was 109, relative to lean mass units (mL/lean-kg/min) was 1.7 and relative to total body mass units (mL/kg/min) was 1.7. Pre-post CR total body mass reduction was −1.3 (3.2) kg and lean body mass gain was 0.8±2.9 kg. BMI similarly had a delta of −0.4 (kg/m^2^). All changes were statistically significant (p<0.05). Results are shown in Table 2.

**Table 2:** Pre-Post cardiac rehabilitation (CR) metrics are shown for cardiopulmonary exercise stress tests (CPET) and dual energy X-ray absorptiometry (DXA) as means (SD) of percent (%) change and absolute delta. *p-values <0.05 represent statistical significance. VO_2_ peak = peak oxygen uptake; BMI = body mass index.

| Pre-Post CR CPET and DXA Metrics |  |  |  |  |  |
| --- | --- | --- | --- | --- | --- |
| Metric | Pre-CR | Post-CR | % Change | Delta | p-value (95% CI) |
| VO<sub>2</sub> Peak (mL/min) | 2,014 (675) | 2,123 (699) | 6.5 (13.1) | 1.1 | <0.001 (64.9, 152)* |
| VO<sub>2</sub> Peak (mL/kg/min) | 22.2 (6.0) | 23.9 (6.5) | 8.2 (13.6) | 1.7 | <0.001 (1.2, 2.2)* |
| VO<sub>2</sub> Peak (mL/lean-kg/min) | 36.1 (8.2) | 37.8 (8.3) | 5.7 (13.4) | 1.7 | <0.001 (1.0, 2.5)* |
| Total Mass (kg) | 91.0 (20.8) | 89.7 (20.2) | -1.3 (3.2) | -1.3 | <0.001 (-1.8, -0.8)* |
| Lean Mass (kg) | 55.3 (10.7) | 55.7 (10.5) | 0.8 (2.9) | 0.40 | 0.005 (0.1, 0.7)* |
| BMI (kg/m<sup>2</sup>) | 30.2 (5.8) | 29.8 (5.7) | -0.1 (1.8) | -0.4 | <0.001 (-0.6, -0.2)* |

Most patients achieved improvements in VO_2_ peak following CR with a fraction of non-responders. Waterfall and bar plots of data are shown in Figure 1. The post-CR percent of patients with a positive change in VO_2_ peak was 70.0%, 75.0%, and 67.9% in the grouped units of ml/min, ml/kg/min, ml/lean-kg/min, respectively. These are shown in Figure 2.

**Figure 1:**
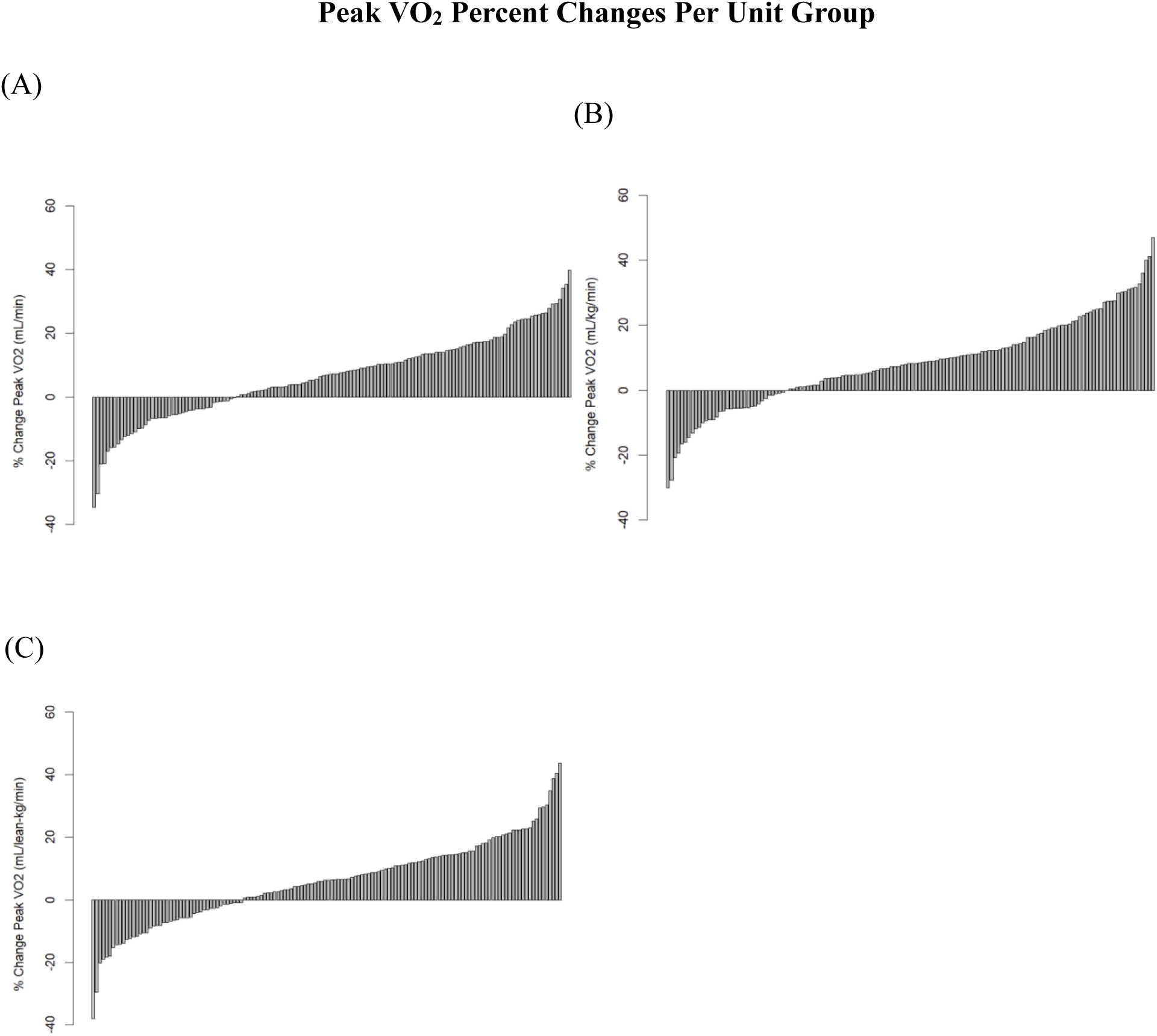
Percent changes of peak oxygen uptake (VO_2_ peak) are plotted by respective units. **(A)** Absolute VO_2_ peak in ml/min **(B)** Relative to total body mass VO_2_ peak in ml/kg/min **(C)** Relative to lean body mass VO_2_ peak in ml/lean-mass/min **(D)** Proportion of patients by percentage having a positive percent change of VO_2_ peak by grouped units.

**Figure 2:**
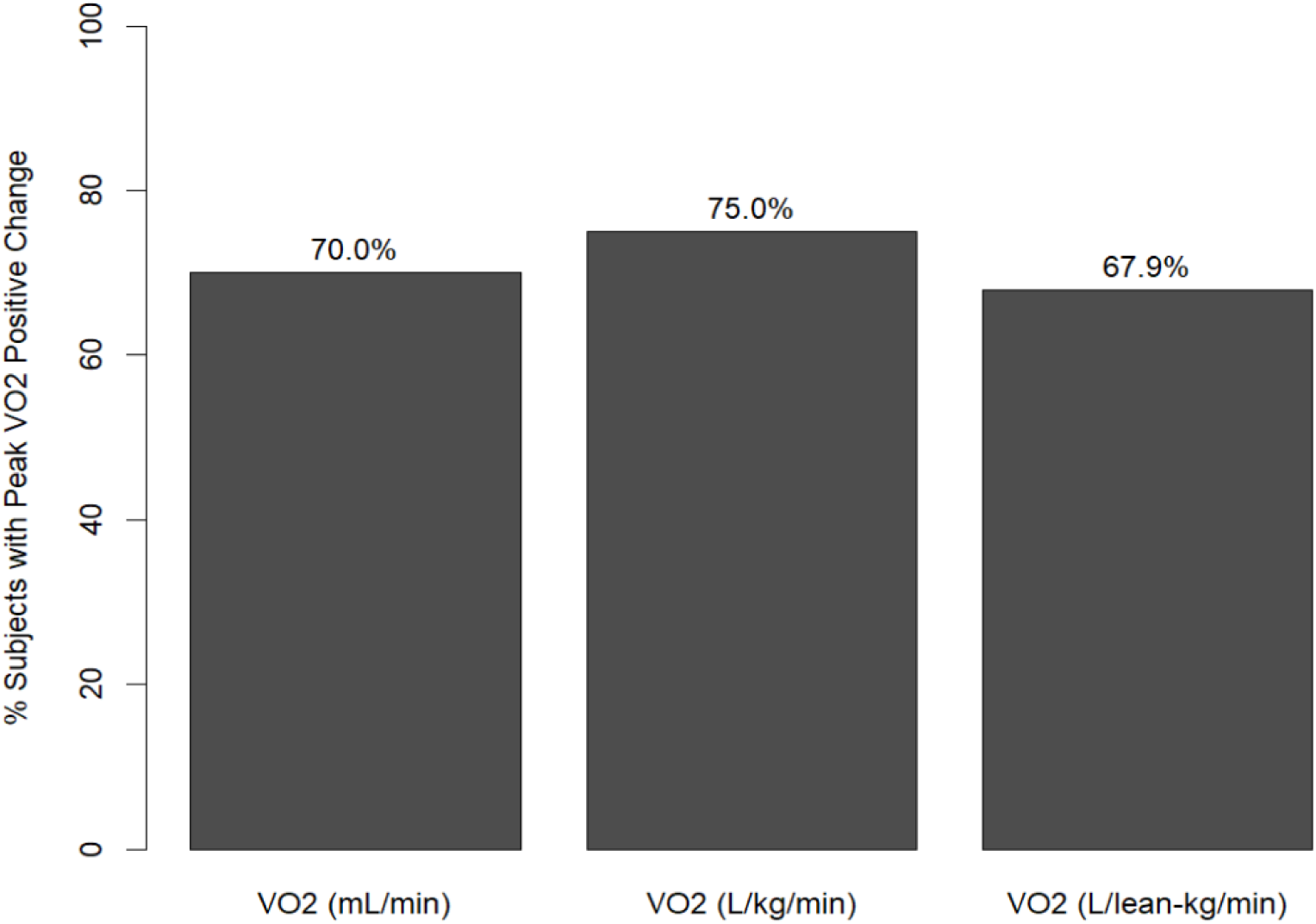
Proportion of patients by percentage having a positive percent change of peak oxygen uptake (VO_2_ peak) by grouped units.

Univariate and multivariate linear regression analyses were completed for VO_2_ peak percent change using several predictors and stratified by unit group. Models were explored for all listed predictors shown in Table 3. These included age, female sex, number of CR sessions, pre-CR BMI, and pre-CR VO_2_. Assuming all else fixed, patients with higher pre-CR VO_2_ peak across all unit groups experienced lower post-CR VO_2_ peak mean percent change per unit increase of pre-CR VO_2_ peak [−0.01 (−0.08, −0.01) ml/min, −0.45 (−0.81, −0.10) ml/kg/min, −0.48 (−0.74, −0.23) ml/lean-kg/min]. Of the tested univariate predictors, only pre-CR VO_2_ for all unit groups showed statistical significance (p <0.05). After adjusting for these predictors in the multivariate model, there was found to be statistical significance only for female sex for absolute VO_2_ units [−7.12 (−13.5, −0.74) ml/min] and pre-CR VO_2_ [−0.01 (−0.02, −0.01) ml/min, −0.73 (−1.17, −0.28) ml/kg/min, −0.59 (−0.89, −0.30) ml/lean-kg/min] for all VO_2_ unit groups. The remaining predictors revealed varying directions and magnitude of change without statistical significance.

**Table 3:** Univariable and multivariable linear regression model outputs for cardiac rehab (CR) patients stratified by peak oxygen uptake (VO_2_ peak) per unit group are shown, including beta coefficients with standard errors (SE), 95% confidence intervals (CI), and p-values. *p-values <0.05 represent statistical significance.

| Variable Linear Regression Model Outputs |  |  |  |  |  |  |
| --- | --- | --- | --- | --- | --- | --- |
| Predictors | Univariable |  |  | Multivariable |  |  |
|  | Beta (SE) | CI (95%) | p-value | Beta (SE) | CI (95%) | p-value |
| VO<sub>2</sub> peak (ml/min) |  |  |  |  |  |  |
| Age | 0.03 (0.09) | (-0.14, 0.21) | 0.720 | -0.10 (0.09) | (-0.29, 0.08) | 0.275 |
| Female Sex | -0.82 (2.81) | (-6.39, 4.74) | 0.770 | -7.12 (3.23)* | (-13.5, -0.74)* | 0.029* |
| Number CR Sessions | 0.02 (0.09) | (-0.17, 0.20) | 0.863 | -0.07 (0.10) | (-0.26, 0.12) | 0.479 |
| Pre-CR BMI | -0.19 (0.19) | (-0.57, 0.19) | 0.327 | 0.20 (0.20) | (-0.22, 0.62) | 0.357 |
| Pre-CR VO<sub>2</sub> | -0.01 (0.00)* | (-0.08, -0.01)* | 0.002* | -0.01 (0.00)* | (-0.02, -0.01)* | <0.001* |
| VO<sub>2</sub> peak (ml/kg/min) |  |  |  |  |  |  |
| Age | 0.00 (0.09) | (-0.18, 0.18) | 0.997 | -0.10 (0.10) | (-0.30, 0.09) | 0.290 |
| Female Sex | -0.19 (2.93) | (-5.98, 5.61) | 0.950 | -3.65 (3.16) | (-9.91, 2.61) | 0.250 |
| Number CR Sessions | -0.01 (0.10) | (-0.57, 0.19) | 0.912 | -0.07 (0.10) | (-0.27, 0.13) | 0.475 |
| Pre-CR<br>BMI | -0.13 (0.20) | (-0.53, 0.26) | 0.505 | -0.35 (0.21) | (-0.77, 0.07) | 0.099 |
| Pre-CR<br>VO<sub>2</sub> | -0.45 (0.18)* | (-0.81, -0.10)* | 0.014* | -0.73 (0.23)* | (-1.17, -0.28)* | 0.002* |
| **VO<sub>2</sub> peak (ml/lean-kg/min)** |  |  |  |  |  |  |
| Age | 0.10 (0.09) | (-0.08, 0.28) | 0.295 | 0.03 (0.10) | (-0.22, 0.16) | 0.746 |
| Female Sex | -1.45 (2.88) | (-7.15, 4.25) | 0.616 | -3.15 (2.87) | (-8.82, 2.52) | 0.274 |
| Number CR<br>Sessions | 0.02 (0.10) | (-0.17, 0.22) | 0.808 | -0.06 (0.10) | (-0.26, 0.13) | 0.516 |
| Pre-CR<br>BMI | -0.10 (0.20) | (-0.49, 0.29) | 0.607 | -0.04 (0.20) | (-0.43, 0.34) | 0.832 |
| Pre-CR<br>VO<sub>2</sub> | -0.48 (0.13)* | (-0.74, -0.23)* | <0.001* | -0.59 (0.15)* | (-0.89, -0.30)* | <0.001* |

Interaction terms were not considered with the limited number of significant predictors in our multivariable model.

## DISCUSSION

The purpose of this study was to assess the influence of body composition on change in VO_2_ following CR. The primary findings of this study are: 1) Regardless of units used for describing change in cardiorespiratory fitness, approximately 30% of patients continue to demonstrate no improvement in VO_2_ peak, 2) The magnitude of change in VO_2_ peak is partly dependent on units used to describe the metric, resulting in an approximate 7% swing in the number of patients who demonstrate improvement in VO_2_ based only on the united used to report VO_2_, 3) Regardless of units used to describe the outcome, similar predictors surface for predicting improvement in VO_2_.

The gold standard metric for assessment of cardiorespiratory fitness remains the measurement of VO_2_ peak during a cardiopulmonary exercise test (CPET) [1]. The units used to describe VO_2_ peak are often made relative to an individual’s body mass to provide comparable units across the spectrum of different sized individuals. For most baseline or static measurements, this works well; however, when individuals are engaged in an intervention that could impact both VO_2_ and body mass, the ability to determine improvement in cardiorespiratory fitness relative to the change in body mass is complex. Further complicating this relationship is the change in body composition for which the total body mass is made up. Reductions in fat mass with increases in metabolically dominant lean mass may result in no change to total body mass while having a significantly positive impact on body composition and overall metabolic health [13]. Concomitant lifestyle changes also play a role. These intricate relationships have not been fully elucidated in patients enrolled in CR, in which both cardiorespiratory fitness and body composition are meaningful outcomes.

Our data confirms that such changes in body mass composition impact reporting of oxygen consumption. Pre-post CR VO_2_ peak relative to total body mass units (mL/kg/min) had the largest statistically significant percent change increase followed by absolute units (mL/min), and relative to lean mass units (mL/lean-kg/min). However, the pre-post CR delta was significantly highest for VO_2_ peak in absolute units (mL/min), followed by units relative to lean mass units (mL/lean-kg/min) and lastly relative to total body mass units (mL/kg/min). The larger reduction in total body mass relative to improvement in lean mass may partly explain why reporting VO_2_ peak in units relative to lean body mass demonstrated a smaller percent improvement relative to other reporting units. It is also worthwhile to consider that since fat mass consumes an exceptionally small amount of oxygen, greater non-lean mass reductions relative to lean mass increases may lend an observed rise in the traditional VO_2_ peak relative to total mass (ml/kg/min) in the absence of an actual rise in peak aerobic capacity.

After normalizing by body mass composition and adjusting for predictors of age, number of CR sessions, pre-CR BMI, and pre-CR VO_2_ in the multivariable regression models, pre-CR VO_2_ for all VO_2_ unit groups was found statistically significant. Patients with higher pre-CR VO_2_ peak experienced lower mean percent change post-CR VO_2_ peak. Lower pre-CR VO_2_ being a significant predictor for greater improvements in post-CR VO_2_, regardless of reported units, is consistent with some recent studies the literature [3, 14].

Our data highlights that the units used to report VO_2_ peak influences the magnitude of change seen following CR. Total body mass compared to lean body mass standardization reflected the greatest percent change improvement, but lean mass standardization reflected the greatest mean delta. Measuring VO_2_ peak using units traditionally relative to total body mass may not necessarily be better than units relative to lean body mass knowing the variable changes in fat mass and lean mass with CR. The beneficial effects of CR on VO_2_ peak, independent of other predictors and despite the modest increases, support this metric remaining a primary goal for CR enrollment given its established relationship to improved mortality [1, 15].

Additionally, between 25% to 32.1% of our patient cohort across all VO_2_ peak unit groups had a seemingly poor response to CR. It has been previously described that approximately 20-30% of patients completing CR could be termed “non-responders” who may reflect neutral or decreased change in VO_2_ peak [3]. VO_2_ peak changes post-CR can also vary widely in the literature [3, 16]. The exact mechanisms remain unclear, though the ability to increase oxygen uptake following CR may potentially outpace the increase in lean body mass to subsequently impart an overall improvement in VO_2_ peak relative to lean body mass. Evidence in young athletes suggests that the initial phase of a new exercise training program results primarily in neuromuscular facilitation changes with minimal change in lean body mass or mitochondrial function [17, 18]. Afterwards, lean mass and mitochondrial changes become exercise intensity, volume, and dose dependent.

Our findings could imply that a proportion of patients continue to demonstrate additional abnormalities (e.g., comorbidities) that inhibit improvement in VO_2_ peak independent of change in body composition. This may partially explain why a subset of patients do not have a positive percent change in VO_2_ peak. Despite CR significantly improving lean body mass, the ability to utilize that mass to generate useful oxidative metabolism could be limited and may require a longer duration of CR to maximize oxidative metabolic efficiency. The oxidative metabolic efficiency of newly acquired lean mass may therefore require a longer duration of CR to maximize expected individual fitness improvements. Given exercise intensity may play a significant role, incorporating high-intensity interval training into CR programs may prove beneficial [19, 20, 21].

Exercise intensity, patient adherence, CR indication, age, and several other factors also play a role. Exercise-based CR has been consistently shown to lead to meaningful improvements in VO_2_ peak for certain patient subsets [22, 23]. Interestingly, however, some recent studies including a clinical trial and systematic review have even challenged the notion that CR imparts significant improvements in measures like VO_2_ peak, mortality, or fitness [24, 25]. Potential reasons include insufficient exercise duration or intensity, patient compliance with the program, or a lack of sufficient statistical power imparted in interventional studies [21]. Improved medical management for patients suffering from cardiovascular events may overshadow benefits from CR previously described in the literature as well.

Cardiac rehabilitation programs were originally designed with exercise training as the primary intervention, designed to improve functional capacity and elicit the multitude of health benefits associated with regular structured exercise. Additionally, counselling on general physical activity is included as a corollary to exercise training and plays an important part in minimization of sedentary behavior. Further, as obesity is a major risk factor for the development and progression of cardiovascular disease, education and counselling focused on nutrition and dietary change to meaningfully improve behaviors associated with poor nutritional choices are now standard of practice for CR programs. These interventions work together to improve cardiorespiratory fitness, improve body composition, and engender positive behavior change for CR patients.

This study examined the impact of body composition on change in VO_2_ per unit group in CR patients. However, we acknowledge potential limitations. The population included in this study arises from a single center with a fairly homogenous population. Our sample was predominately male and Caucasian potentially limiting generalizability based on demographics. Further, we only included individuals who had completed both pre- and post-CR measurements of VO_2_ peak as well as body composition resulting in a limited sample size. Additionally, this is a retrospective cohort study and by design inference of causality cannot be made.

## CONCLUSION

Significant decreases in total body mass and increases in lean body mass and VO_2_ occur following CR regardless of units reported, seeming to contribute a meaningful portion of peak VO_2_ improvements. Larger improvements are also reflected in units adjusted for total mass compared to other units. Lower pre-CR VO_2_ is a significant predictor for greater improvements in post-CR VO_2_. These data suggest the reporting methodology for change in VO_2_ following CR may impact overall findings.

## Data Availability

Data are to be made available upon reasonable request.

## ACKNOWLEDMENTS

The authors wish to acknowledge the dedicated nursing and cardiology rehabilitation staff who contributed to the care of our patients.

## REFERENCES

1. Mandsager K, Harb S, Cremer P, Phelan D, Nissen SE, Jaber W. Association of Cardiorespiratory Fitness with Long-term Mortality Among Adults Undergoing Exercise Treadmill Testing. JAMA Netw Open. 2018;1(6):e183605. doi:10.1001/jamanetworkopen.2018.3605

2. Simon M, Korn K, Cho L, Blackburn GG, Raymond C. Cardiac rehabilitation: A class 1 recommendation. Cleve Clin J Med. 2018 Jul;85(7):551–558. doi: 10.3949/ccjm.85a.17037. PMID: 30004380.

3. Little KA, Smith JR, Medina-Inojosa JR, Chacin Suarez AS, Taylor JL, Hammer SM, Fischer KM, Bonikowske AR, Squires RW, Thomas RJ, Olson TP. Predictors of Changes in Peak Oxygen Uptake After Outpatient Cardiac Rehabilitation: Importance of Cardiac Rehabilitation Attendance. Mayo Clin Proc Innov Qual Outcomes. 2022 Sep 2;6(5):428–435. doi: 10.1016/j.mayocpiqo.2022.07.002. PMID: 36097546; PMCID: PMC9463170.

4. Krachler B, Savonen K, Komulainen P, Hassinen M, Lakka TA, Rauramaa R. Cardiopulmonary fitness is a function of lean mass, not total body weight: The DR’s EXTRA study. Eur J Prev Cardiol. 2015 Sep;22(9):1171–9. doi: 10.1177/2047487314557962. Epub 2014 Nov 7. PMID: 25381337.

5. Martin A, Fox D, Murphy CA, Hofmann H, Koehler K. Tissue losses and metabolic adaptations both contribute to the reduction in resting metabolic rate following weight loss. Int J Obes (Lond). 2022 Jun;46(6):1168–1175. doi: 10.1038/s41366-022-01090-7. Epub 2022 Feb 18. PMID: 35181758; PMCID: PMC9151388.

6. Mondal H, Mishra SP. Effect of BMI, Body Fat Percentage and Fat Free Mass on Maximal Oxygen Consumption in Healthy Young Adults. J Clin Diagn Res. 2017 Jun;11(6):CC17–CC20. doi: 10.7860/JCDR/2017/25465.10039. Epub 2017 Jun 1. PMID: 28764152; PMCID: PMC5535345.

7. Maciejczyk M, Więcek M, Szymura J, Szyguła Z, Wiecha S, Cempla J. The influence of increased body fat or lean body mass on aerobic performance. PLoS One. 2014 Apr 21;9(4):e95797. doi: 10.1371/journal.pone.0095797. PMID: 24752377; PMCID: PMC3994154.

8. von Elm E, Altman DG, Egger M, Pocock SJ, Gøtzsche PC, Vandenbroucke JP; STROBE Initiative. Strengthening the Reporting of Observational Studies in Epidemiology (STROBE) statement: guidelines for reporting observational studies. Lancet. 2007 Oct 20;370(9596):1453-7. doi: 10.1016/S0140-6736(07)61602-X. PMID: 18064739.

9. Thomas RJ, King M, Lui K, Oldridge N, Piña IL, Spertus J; ACC/AHA Task Force Members. AACVPR/ACC/AHA 2007 performance measures on cardiac rehabilitation for referral to and delivery of cardiac rehabilitation/secondary prevention services. J Cardiopulm Rehabil Prev. 2007 Sep-Oct;27(5):260–90. doi: 10.1097/01.HCR.0000291295.24776.7b. PMID: 17885506.

10. Noble BJ. Clinical applications of perceived exertion. Med Sci Sports Exerc. 1982;14(5):406–11. doi: 10.1249/00005768-198205000-00016. PMID: 7154897.

11. Squires RW, Allison TG, Johnson BD, Gau GT. Non-physician supervision of cardiopulmonary exercise testing in chronic heart failure: safety and results of a preliminary investigation. J Cardiopulm Rehabil. 1999 Jul-Aug;19(4):249–53. doi: 10.1097/00008483-199907000-00006. PMID: 10453432.

12. Guazzi M, Arena R, Halle M, Piepoli MF, Myers J, Lavie CJ. 2016 focused update: clinical recommendations for cardiopulmonary exercise testing data assessment in specific patient populations. Eur Heart J. 2018 Apr 7;39(14):1144–1161. doi: 10.1093/eurheartj/ehw180. PMID: 27141094.

13. Brown TM, Pack QR, Aberegg E, Brewer LC, Ford YR, Forman DE, Gathright EC, Khadanga S, Ozemek C, Thomas RJ; American Heart Association Exercise, Cardiac Rehabilitation and Secondary Prevention Committee of the Council on Clinical Cardiology; Council on Cardiovascular and Stroke Nursing; Council on Lifestyle and Cardiometabolic Health; and Council on Quality of Care and Outcomes Research. Core Components of Cardiac Rehabilitation Programs: 2024 Update: A Scientific Statement From the American Heart Association and the American Association of Cardiovascular and Pulmonary Rehabilitation. Circulation. 2024 Oct 29;150(18):e328–e347. doi: 10.1161/CIR.0000000000001289. Epub 2024 Sep 24. Erratum in: Circulation. 2025 Apr 29;151(17):e965-e966. doi: 10.1161/CIR.0000000000001338. PMID: 39315436.

14. Eduardo, L, Ritt, F, Rocco, E. et al. Low VO2 Peak Is Independently Associated with Improvement in Functional Capacity After Cardiac Rehabilitation: Results from a National and Multicenter Study. JACC. 2018 Mar, 71 (11_Supplement) A1885. 10.1016/S0735-1097(18)32426-4.

15. Rouleau CR, Chirico D, Wilton SB, MacDonald MK, Tao T, Arena R, Campbell T, Aggarwal S. Mortality Benefits of Cardiac Rehabilitation in Coronary Artery Disease Are Mediated by Comprehensive Risk Factor Modification: A Retrospective Cohort Study. J Am Heart Assoc. 2024 May 21;13(10):e033568. doi: 10.1161/JAHA.123.033568. Epub 2024 May 18. PMID: 38761079; PMCID: PMC11179828.

16. Carbone S, Kim Y, Kachur S, Billingsley H, Kenyon J, De Schutter A, Milani RV, Lavie CJ. Peak oxygen consumption achieved at the end of cardiac rehabilitation predicts long-term survival in patients with coronary heart disease. Eur Heart J Qual Care Clin Outcomes. 2022 May 5;8(3):361–367. doi: 10.1093/ehjqcco/qcab032. PMID: 33895797; PMCID: PMC9631231.

17. Hong AR, Hong SM, Shin YA. Effects of resistance training on muscle strength, endurance, and motor unit according to ciliary neurotrophic factor polymorphism in male college students. J Sports Sci Med. 2014 Sep 1;13(3):680–8. PMID: 25177199; PMCID: PMC4126309.

18. Huertas JR, Casuso RA, Agustín PH, Cogliati S. Stay Fit, Stay Young: Mitochondria in Movement: The Role of Exercise in the New Mitochondrial Paradigm. Oxid Med Cell Longev. 2019 Jun 19;2019:7058350. doi: 10.1155/2019/7058350. Erratum in: Oxid Med Cell Longev. 2021 Jan 18;2021:9274841. doi: 10.1155/2021/9274841. PMID: 31320983; PMCID: PMC6607712.

19. McGregor G, Powell R, Begg B, Birkett ST, Nichols S, Ennis S, McGuire S, Prosser J, Fiassam O, Hee SW, Hamborg T, Banerjee P, Hartfiel N, Charles JM, Edwards RT, Drane A, Ali D, Osman F, He H, Lachlan T, Haykowsky MJ, Ingle L, Shave R. High-intensity interval training in cardiac rehabilitation: a multi-centre randomized controlled trial. Eur J Prev Cardiol. 2023 Jul 12;30(9):745–755. doi: 10.1093/eurjpc/zwad039. PMID: 36753063.

20. Dun Y, Smith JR, Liu S, Olson TP. High-Intensity Interval Training in Cardiac Rehabilitation. Clin Geriatr Med. 2019 Nov;35(4):469–487. doi: 10.1016/j.cger.2019.07.011. Epub 2019 Jul 12. PMID: 31543179; PMCID: PMC6760312.

21. Taylor JL, Bonikowske AR, Olson TP. Optimizing Outcomes in Cardiac Rehabilitation: The Importance of Exercise Intensity. Front Cardiovasc Med. 2021 Sep 3;8:734278. doi: 10.3389/fcvm.2021.734278. PMID: 34540924; PMCID: PMC8446279.

22. Sheng SP, Feinberg JL, Bostrom JA, Tang Y, Sweeney G, Pierre A, Katz ES, Whiteson JH, Haas F, Dodson JA, Halpern DG. Adherence and Exercise Capacity Improvements of Patients With Adult Congenital Heart Disease Participating in Cardiac Rehabilitation. J Am Heart Assoc. 2022 Aug 16;11(16):e023896. doi: 10.1161/JAHA.121.023896. Epub 2022 Aug 5. PMID: 35929458; PMCID: PMC9496295.

23. Tegegne TK, Rawstorn JC, Nourse RA, Kibret KT, Ahmed KY, Maddison R. Effects of exercise-based cardiac rehabilitation delivery modes on exercise capacity and health-related quality of life in heart failure: a systematic review and network meta-analysis. Open Heart. 2022 Jun;9(1):e001949. doi: 10.1136/openhrt-2021-001949. PMID: 35680170; PMCID: PMC9185675.

24. Nichols S, Taylor C, Goodman T, Page R, Kallvikbacka-Bennett A, Nation F, Clark AL, Birkett ST, Carroll S, Ingle L. Routine exercise-based cardiac rehabilitation does not increase aerobic fitness: A CARE CR study. Int J Cardiol. 2020 Apr 15;305:25–34. doi: 10.1016/j.ijcard.2020.01.044. Epub 2020 Jan 22. Erratum in: Int J Cardiol. 2021 Jan 1;322:294. doi: 10.1016/j.ijcard.2020.08.078. PMID: 32008847.

25. Sandercock GR, Cardoso F, Almodhy M, Pepera G. Cardiorespiratory fitness changes in patients receiving comprehensive outpatient cardiac rehabilitation in the UK: a multicentre study. Heart. 2013 Jun;99(11):785–90. doi: 10.1136/heartjnl-2012-303055. Epub 2012 Nov 24. PMID: 23178183.

